# Uptake of Covid-19 preventive measures among 10 immigrant ethnic groups in Norway

**DOI:** 10.1101/2021.11.24.21266682

**Authors:** Abdi Gele, Naima Said Sheikh, Prabhjot Kour, Samera A Qureshi

## Abstract

**Background:** A pessimistic view of the impact of Covid-19 on immigrants has generated an interest in exploring the role of socio-economic and cultural factors on excess infection, hospitalization and death among immigrants. Nowhere in the world is such interest more palpable than in Western countries, including Norway. An expanding amount of literature has demonstrated that preexisting socio-economic inequalities have affected Covid-19 control programs through a disruption of immigrants’ uptake to preventive measures. Nonetheless, until very recently, no qualitative research has been conducted to address the impact of socio-economic and socio-cultural factors on immigrants’ uptake on preventive measures of Covid-19 in Norway.

**Methods:** An interview-based qualitative study consisting of 88 participants (49 women and 39 men) from 10 immigrant ethnic groups were carried out. Participants were recruited through purposive sampling and snowballing. In-depth interviews were held through telephone or online for those who have experience in the use of zoom or teams. Data were analyzed using thematic analysis

**Results:** We found that participants’ attitudes toward the pandemic in general, and more specifically their adherence to preventive measures, have increased over time. However, the number of barriers that hinder immigrants from adhering to preventive measures were identified and classified more broadly into three main subthemes: 1) socio-economic barriers; 2) socio-cultural barriers, and 3) other barriers. Socio-economic barriers include overcrowded households, working in first-line jobs, education and language. Socio-cultural barriers include collectivist culture, religious fatalism and risk perception toward the pandemic.

**Conclusion:** To reduce the health inequality that arises from overcrowded housing, there is a need for a long-term strategy to help improve the housing situation of low-income immigrant families that live in overcrowded households. In addition, increasing health literacy and more generally, the integration of immigrants, may also reduce the effect of socio-cultural factors on an immigrant’s uptake of preventive measures.

## Background

A novel coronavirus known as SARS-CoV-2 emerged in China in late 2019, thereby causing the disease known as Covid-19. The World Health Organization declared Covid-19 to be a pandemic on March 12, 2020; since that time the pandemic has been met by unequal responses among countries, and led to unequal impacts on communities. Most global nations considered non-chemotherapeutic measures such as wearing face masks, maintaining a social distance from other people, lockdowns, avoiding large gatherings and hygiene practices to be the key tools in slowing the spread of Covid-19 (1). The success of those measures largely relies on the capacity, willingness and motivation of the population to comply with them. A prior study found that adherence to mitigation measures is followed by a significantly lower mortality from Covid-19 at the community level (2). The factors that may determine people’s adherence to mitigation measures include the form and perception of the governmental communication, as well as socio-cultural and socio-economic factors (2). A recent review also found that the perceived risk and knowledge of the disease, knowledge about- and the perceived benefits of quarantine, social norms and financial consequences all affect adherence (3).

Although Covid-19 poses a danger to everyone, there is evidence that it is taking a particularly heavy toll on immigrant populations (4, 5). In Scandinavian countries, the Covid-19 pandemic has exposed preexisting social and health disparities, with stark differences in the proportion of immigrants and non-immigrants who have been diagnosed, hospitalized and died from Covid-19 (6-8). This raises concern about the role of pervasive social determinants that have long adversely impacted the health of immigrants. Predisposing factors such as living and working conditions (9), cultural and language barriers, poor knowledge and limited social networks (10) may limit the ability of migrant populations to access adequate health care and eventually avoid infection (11). Literature suggests that Covid-19 response measures should address underlying conditions of vulnerability linked with migrants’ socio-economic situation (12).

A study in Norway found a notification and hospitalization rate of 251, 21 per 100,000 respectively, for non-immigrants, and 567 and 61 per 100,000, respectively, for immigrants. The notification rate was highest among immigrants from Somalia (2057), Pakistan (1868), Iraq (1616) and Afghanistan (1391) (6). Factors such as crowded housing and low income at a group level were correlated with rates of both notified cases of Covid-19 (Pearson’s correlation coefficient 0.77 and 0.52) and related hospitalizations (0.72, 0.50) among immigrants in Norway (13). The same study reported that a low educational level and unemployment were correlated with a high number of notified cases. However, a register data analysis shows that the excess risk of immigrants to the infection does not seem to be fully explained by differences in socioeconomic status (14). A lack of awareness of locally recommended prevention measures, an overreliance on informal communication channels or an adherence to culture-specific customs may also partially explain these differences. Thus far, most of the studies on Covid-19 among immigrants in Norway have been based on a register data analysis. Hence, there is a need for qualitative studies that explore immigrants’ perspectives of why they are disproportionately affected by Covid-19, in addition to factors that hinder them from adhering to mitigation measures. This paper explores immigrants’ perspectives of the factors that impede their adherence to preventive measures against Covid-19, which may also explain their overrepresentation on Covid-19 infection in Norway.

## Methods

### Recruitment

The sample consisted of members who were recruited from a resource group that served as a breach between community and service providers. Others were recruited from researchers’ networks and through snowballing. Researchers from the Norwegian Institute of Public Health (NIPH), and the research consultancy firm known as Opinion, recruited the participants using purposive and snowballing methods. We included highly diverse individuals in terms of occupation, education, age and length of residency in Norway. We recruited 88 participants from 10 different ethnic groups in this study: Somalia (12), Iraq (12), Pakistan (12), Afghanistan (11), Poland (11), Sri Lanka (6), Turkey (6), Bosnia/Serbia (6), Eritrea (6) and Syria (6).

### Data collection

To help address the aim of this paper, the coordinating author (AG) identified and engaged with seven multi-disciplinary and multi-lingual researchers from the NIPH, which carried out 34 interviews, while Opinion conducted 54 interviews. We used a predesigned question guide for the interviews specifically developed for this study and reviewed by number of researchers in the field of migration and health. Some immigrants were interviewed in their native languages, whereas others were interviewed in Norwegian or English based on their preference. The interviews lasted from 30 to 90 minutes, and were tape recorded and transcribed verbatim by the interviewers. The study was funded by the Ministry of Education and Integration, Norway.

### Data analyses

The data were transcribed by interviewers, while the transcripts were analyzed using NVivo version 12 by NS and AG. The research objectives formed the basis of the coding, and the analysis of the raw data was completed following an inductive approach, based on a thematic analysis (15). NS and AG carefully read the transcripts several times for accuracy and completeness. The coded excerpts and quotations were reviewed to help understand the link between different concepts, and eventually develop core categories. We later used the constant comparison method and selective coding to identify emergent themes both within and across interviews. We used a thematic analysis to identify and analyze important categories and themes (15). This allowed the research findings (major themes) to emerge naturally from the participants’ interviews, without the restrictions that might be created by more structured methodologies (16). The authors identified recurrent and important themes, and subsequently summarized them under thematic headings. In line with prior studies, we ensured the validity of the themes by repeatedly verifying them against the raw data from which they were originated (17). A cooperative author reflection on the raw data in the group synthesis further verified the reliability of the themes.

## Results

Of the 88 participants, 49 were females and 39 were males, with their ages varying from 19 to 78 years old. Although the majority of the participants lived in Norway for 15 years, some were newly arrived while others were born in Norway. Of those who reported their education level, 30 had a university education, whereas others had a secondary school or lower. As shown in Figure 1, two major themes, four subthemes and 12 categories emerged from the interviews (Figure 1).

**Figure 1:**
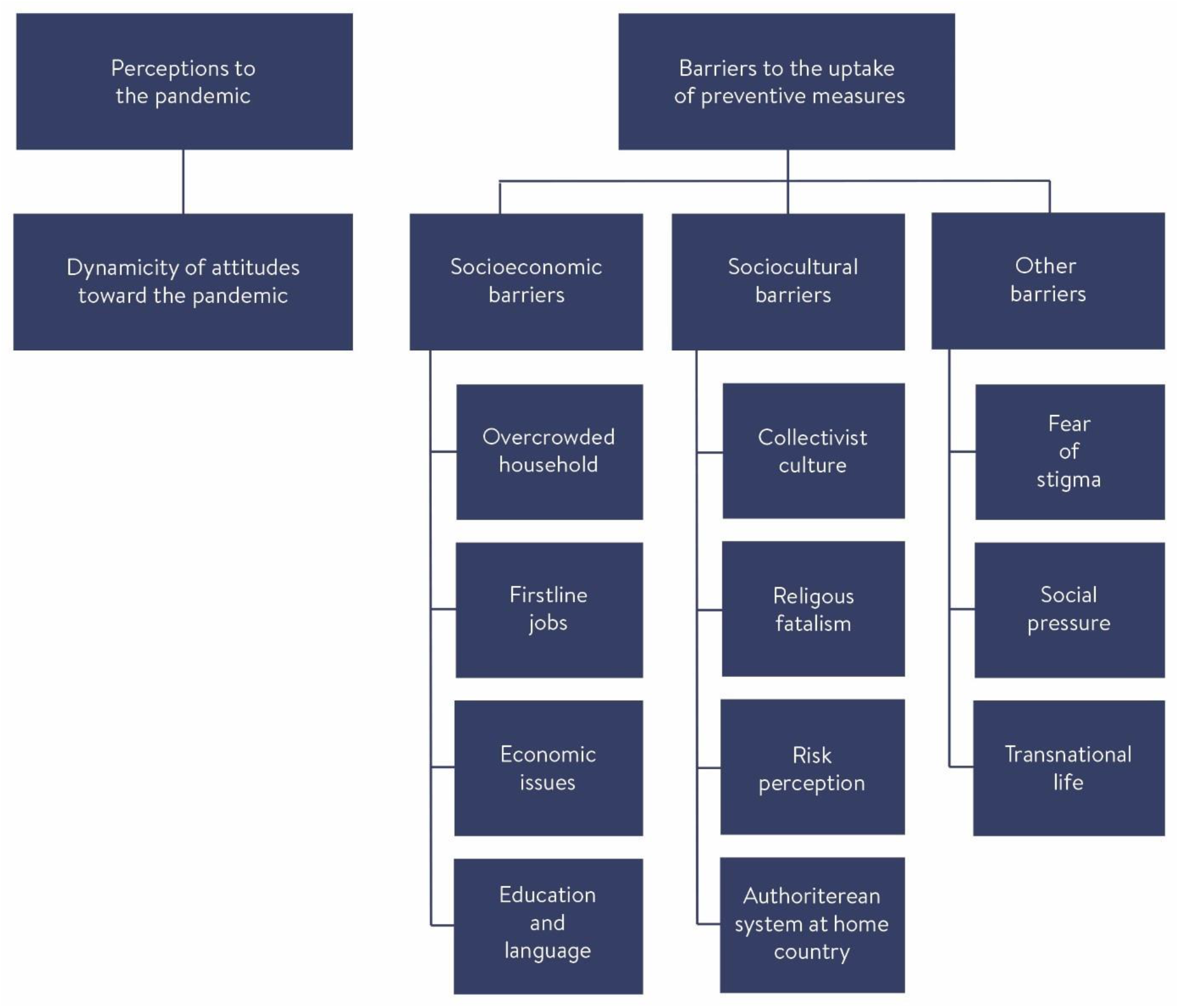
Themes, subthemes and categories.

### Theme 1: Perception toward the pandemic

#### Dynamicity of the perception and knowledge toward the pandemic

According to the study participants, the pandemic has disproportionately affected some immigrant groups in Norway at the beginning of the pandemic. Nonetheless, participants stated that their perceptions and knowledge toward the pandemic have evolved over time. All the different ethnic groups agreed that immigrants’ physical and psychological meaning of living with the pandemic, as well as their motivation in complying with government instructions, have improved:

> *We had a period of unclarity of what to do, how it [the coronavirus] is transmitted, how can we protect ourselves. But now things are getting better we have better knowledge, and we follow regulations of wearing masks, washing hands*. (Iraqi Norwegian male in his 50s)
>
> *People have changed a lot. First, we believed that the virus is not dangerous. Some of us believed that the virus is for Chinese, while other believed that it is not for Muslims. But now we can see that people’s attitudes have changed. Last year (2020), there were celebrations everywhere such as birthdays, weddings, etc. Nowadays people are very careful, there are no more wedding ceremonies and people do not visit each other. This is because they understood how dangerous the virus can be after many of their family members and acquaintances were infected, others hospitalized, while others have died from Covid-19*…(Somali Norwegian female in her 30s)

### Theme 2: Barriers to uptake of preventive measures

#### Socio-economic barriers

Participants were asked questions related to the overrepresentation of immigrants in people infected by the coronavirus, and those being hospitalized due to Covid-19. Participants overwhelmingly mentioned that overcrowded households and first-line jobs, as well as education and language, are among contributing factors for a higher infection rate among their communities.

#### Overcrowded households

According to several participants, many immigrant families live in housing conditions that make them vulnerable to becoming infected. Almost all ethnic groups agreed that overcrowded households make social distancing very difficult. Some participants stated that once a family member is infected, the chance of passing the infection to other family members is extremely high. They also highlighted that quarantine and isolation are rarely feasible for many immigrant families, as they do not have enough space and rooms to isolate their infected family member:

> *Many immigrants live in overcrowded living conditions. If a member in the family gets infected, the other members in the family have no chance to protect themselves from the virus. I know an Afghan family whose daughter got the infection at work*… *The whole family became infected*. (Afghani Norwegian female in her 50s)
>
> *It has always been difficult for some people to practice distancing and follow the quarantine rules, because of several individuals living in a small apartment. The housing problem is not something you can do anything about it, because people live by their financial capacity* (Somali Norwegian female in her 30s).

Although participants have reported overcrowded households as a major source of Covid-19 transmission in their communities, they also reported extended families visiting each other, thereby making the already overcrowded households even more congested:

> *When I’m at home, things get a little harder because it’s not just me who decides (on whether people visit or not). At times, we receive three visitors. We are already seven people living there, so we become 10 persons together. (*Afghani Norwegian female in her 20s)

#### First-line jobs

Participants from all ethnic groups have indicated that their communities account for most of the first-line workers in Norway, such as care workers, the transport sector, construction and cleaning, etc., in which it is impossible to work from home. It is not only that they have to go to work in those specific sectors; they also have to deal with subsequent unsafe working conditions with respect to Covid-19 transmission:

> *Covid-19 has affected me, my family and my community to a greater extent. I think there are two reasons for this: first, most of the people in my community work first-line jobs; they have frequent contact with different people, for example, taxi/bus drivers, grocery store workers, the healthcare sector, etc. It is impossible to work from home and stay isolated. Their work exposes them to increased risk of Coronavirus infection*. (Pakistani Norwegian male in his 70s)

Participants reported that when immigrant workers are infected with Covid-19 at their workplace, they pass the infection to family members. The immigrant’s vulnerability to the coronavirus transmission at the workplace, accompanied with a large family who live in overcrowded households, may create a circle of transmission among immigrants and make the chain of transmission difficult to break:

> *There are many people who work in first-line jobs such as kindergartens, construction sites, health services and shops, so all my friends became infected at work because they cannot have a home office. They pass the infection to their families* (Polish Norwegian female in her 40s).

#### Financial barrier

Participants stated that there are large families with a low-income who may not be able to afford to cover the cost of buying face masks and sanitizers to everyone in the household. According to one participant, families may prioritize their basic needs, such as food, clothes and medicine for their limited income. Therefore, they may be exposed to the infection, as they cannot adhere to some critical prevention measures such as face masks:

> *We have our own plans and take our decisions regarding expenses. For example, I can’t afford using one-time-use face masks. I’ve a family of six; if the face mask box with 10 pieces costs 150 NOK, and my monthly income is low, I’m using cloth-made face masks, which may not be very effective. We spoke about the social and cultural factors, but there also economic ones here. The face mask is expensive, and it increases my monthly expenses*. (Iraqi Norwegian female in her 30s)
>
> *It is expensive to adhere to measures such as face masks or driving a car. Those who drive their own cars have a minimum exposure to the virus*. (Eritrean Norwegian female in her 20s)

#### Education and language

Participants related low education with immigrants’ poor understanding of the risk posed by Covid-19. Similarly, language was reported to be a key in receiving, understanding and acting upon the information provided by health authorities. According to participants, language problems were more common with elderly people. Both language and education were mentioned as barriers to the uptake of government instructions:

> *Those with a higher education can understand the risk posed by* Covid-19. *They obtain the necessary information as they follow the media. Those with a low education do not understand the information*… *Since most people have low levels of education, it means that they do not understand the severity of the virus. It is difficult for them to understand*. (Somali Norwegian female in her 30s)
>
> *Language barrier is a huge challenge, especially for the older generation. They do not receive the information because it is in Norwegian which they do not understand*. (Pakistani Norwegian female in her 40s)

### Socio-cultural barriers

#### Collectivist culture

According to the participants, immigrants’ social contact patterns are a crucial factor behind the spread of Covid-19 among immigrants. They stated that the benefit of abiding to strict social distancing rules and reducing mobility tends to be harder for some immigrants, who are accustomed to frequent social interactions:

> *In the Eastern culture, social contact is a very important aspect of our daily routine. I can’t spend a day without meeting with my social network. In my view, I would have preferred to get the coronavirus infection rather than being isolated* (Iraqi Norwegian female in her 20s).

Participants from all ethnic groups stated that there are cultural differences between immigrants and non-immigrants. This difference includes the level of social contacts, as well as the physical distance that people keep when interacting with others:

> *There are cultural differences. We shake hands with each other, we meet every day, so it is difficult to change it (participants demonstrated greeting with elbow, etc*.)… *so it is difficult for us*. (Polish Norwegian female in her 40s)
>
> *Somalis are more social people and we come from a tradition where it is normal to meet frequently, so it is difficult for people who have grown up in a context with a high level of social contact to live in isolation*. (Somali Norwegian female in her 30s)

#### Religious fatalism

A prevalent religious fatalism ideology, which refers to the belief that disease prevention is beyond human control, has been reported by some participants to play a critical role on the spread of infection in some immigrant groups. Participants mention that people with a religious fatalism ideology ignore their personal responsibility, and leave everything to God. Participants also mentioned that religious fatalism is a major hindrance to adherence despite some religious leaders making it clear that a fatalism ideology has no basis in any religion. Some participants stated that religious fatalism is the result of a low level of education among community members:

> *Unfortunately, there are people who say “God created us and we will die when the destiny comes. God decides everything. Whether it is a war, a corona or being hit by a car. So, this is something that is unfortunately going on, and something that we should maybe work on*. (Somali Norwegian female in her 30s)
>
> *Traditionally, religious beliefs play an important role; they believe if God wants to protect them, they will be protected*. (Iraqi Norwegian male in his 30s)
>
> *There are many people who say that their death is predetermined, and so is the reason for their death. Everyone will die at a predetermined moment for a reason; if the reason for your death is Covid-19, nobody can change it, so there is no need to worry. This shows the carelessness of some people in the community*. (Pakistani Norwegian male in his 40s)

#### Risk perception

The participants reported that some immigrants have a different risk perception about Covid-19, which may create an environment in which the perceived susceptibility and perceived severity of the disease are both low. Some participants expressed that their communities’ risk perception about the pandemic is determined by the way this community subjectively estimates and feels about the risk. For example, they said that refugees from war-torn countries, such as Afghanistan, Syria, Somalia and Eritrea, might have experienced indescribable atrocities back home, including killings, the bombing of residences and infectious diseases, which kill more instantly than Covid-19. According to the participants, the risk they experienced back home, which is their reference point, is far bigger than the risk Covid-19 poses to them:

> *They perceive death as something normal. They saw the wars in Syria and Iraq, and that people died in front of their eyes, so this* Covid-19 *is incomparable with what they have experienced*. (Iraqi Norwegian female in her 20s)
>
> *Somali people do not see the risk of* Covid-19. *They don’t care. They grew up in war, and it has become normal for them to see people dying*. (Somali Norwegian, female in her 30s)

#### Authoritarian system in their home country

According to the participants, the success of the effort to control the transmission of Covid-19 depends on immigrants’ trust in both the information provided, and in the institutions responsible for the pandemic control endeavor. For some, they came from contexts in which there is no trust between the government and its people. To demonstrate its significance, when governments introduce new measures, they impose enforcement measures, including punishment, arrests and fines to those who fail to comply. As some immigrants get used to an authoritarian system, government instructions are considered as word-of-mouth, and they barely follow them in the absence of enforcement measures:

> *When it comes to compliance, I think people follow the rules in Eritrea. Everyone has to obey government instructions, because a failure to do so will result in severe consequences. Here, there is more freedom, but people have not gotten used to this type of freedom*. (Eritrean Norwegian female in her 20s)
>
> *There is a group within the Polish community that follow more on instructions in Poland than what is happening in Norway. So, they are very much influenced by the information they received from Poland. The rules in Poland are different from here. Especially last year, regarding compliance with quarantine rules, there was a lot of control, you have to have an app, send photos every single day from the living room, the kitchen and so on. Also, the police come to the door almost every day to check that you are actually sitting at home. In Norway, the system is trust-based. For people who do not know this system, it is difficult for them to understand that it’s an individual’s responsibility to follow quarantine rules*. (Polish Norwegian female in her 40s)

In line with previous quotes, study participants indicated that their communities have a limited understanding of the Norwegian system, in which there is strong trust between the government and the people. Participants expressed that the difference in political culture between countries of origin and Norway have affected immigrants’ adherence to governmental instructions:

> *There is a cultural difference between Poland and Norway. In Poland, if there is no enforcement, people do not follow the rules. In Norway, they use the expression “You should”, you should stay at home, for example, yes, but you can go out. Okay, I can go out. In Poland if you are quarantined, soldiers come to your home every day and you have to show yourself in the window to prove that you are at home. Here, everything is based on trust, while in Poland it is control based. So that’s the difference*. (Polish Norwegian female in her 40s)

### Theme 3: Other barriers

#### Fear of stigma

According to the participants, immigrants have been scapegoated for disease transmission in Norway. Some participants stated that immigrants’ overrepresentation of the infection rate have exposed immigrants to accusations of being negligent, thereby being held responsible for having contracted the virus. The fear of stigma has created an environment in which people hide their Covid-19 status, and avoid being tested or diagnosed.

> *I was traveling on a bus and I sneezed. Many people immediately looked at me while those sitting nearby left their seats. This behavior will influence people’s decisions. People may hide their Covid-19 status if they are infected to avoid such negative attention or potential discrimination*. (Pakistani Norwegian male in his 70s)

#### Social pressure

Some participants reported peer pressure with a potential to hinder their adherence to social distancing measures. Occasional visits to family and friends and participation in social events, such as weddings and birthdays, as well as visiting hospitalized individuals, or participation in burial ceremonies, are considered by some immigrant groups as a social obligation. According to the informants, the violation of social rules regardless of the pandemic can lead to social sanctions and exclusion. The consequence of not abiding social demands may be psycho-socially unbearable; therefore, most of the members may succumb to the social pressure by participating in social activities, regardless of the risk for disease transmission:

> *Keeping a distance from people and thinking about one’s health has been very difficult (with Covid-19) because of the culture and tradition of some communities. (If you reject an invitation for a wedding*.*) People can feel that you do not like them despite explaining the reason. Some people even say, “Yes you have got a justification*.*” I think that is very difficult for some communities*. (Afghani Norwegian female in her 50s)

#### Transnational life

Some participants have expressed that the adherence to Covid-19 mitigation measures has been affected by immigrants’ frequent travel movements. According to them, immigrant members frequently cross Norway’s borders back and forth for work, and for family reasons. They mentioned the policy that people who traveled abroad must quarantine in their home for 14 days from the day they returned to Norway. Participants reported that this can be difficult for communities that live in overcrowded households. According to the participants, there are immigrant individuals who returned to Norway with the virus, thereby transmitting the virus to the members of their families and their communities and networks:

> *There were many who had traveled, and they came back with the virus*. (Afghani Norwegian female in her 50s)
>
> *Yes, many Pakistanis also traveled to Pakistan without any necessary reason, and got infected there and brought back the infection; this can also be a factor*. (Pakistani Norwegian female in her 50s)

## Discussion

The study explored immigrants’ perceptions toward the pandemic and their adherence to Covid-19 preventive measures. We found that the understanding of the pandemic and of preventive measures improved over time, as people received tailored information about the pandemic. In line with our findings, a prior study in Norway shows that an adherence to Covid-19 preventive measures has increased among immigrants over time (18). The study hypothesized that adherence may be influenced by the perceived infection risk, or that the population experienced quarantine fatigue and a desire to return to normality (19). Another study in Norway also found that adherence to quarantine/isolation has been low in Norway after the first wave (19), although this study was not specifically for immigrants. The improved perception of COVID-19 among immigrants may be influenced by the high infection rate and the hospitalization of immigrants in the first wave, and the subsequent targeted intervention directed toward immigrants to contain the transmission among immigrants.

Despite the improved perception and subsequent increased adherence, immigrants have faced socio-economic barriers to adherence. Among these barriers is “living in overcrowded households.” Household crowding is a “condition where the number of occupants exceeds the capacity of the dwelling space available, whether measured as rooms, bedrooms or floor area, resulting in adverse physical and mental health outcomes” (20). Several studies have reported a direct association between crowding and adverse health outcomes, such as infectious diseases (21, 22). Overcrowded households make it harder to self-isolate, physical distancing and the quarantining of suspected cases. An analysis carried out in the UK found a strong correlation between Covid-19 death rates and the level of overcrowded households (22). Our study supports the finding of a prior study in Norway that overcrowded households may contribute to immigrants’ high infection rates in Norway (13). Statistics Norway documented that approximately half of immigrants from Somalia and Pakistan live in overcrowded households (23), which is a source of transmissible diseases to the wider community (24). In line with our findings, a recent study found that the risk of infection increased dramatically with household size, ranging from <0.2% for individuals living alone to 5.4% for households accommodating seven persons (25). The high prevalence of overcrowded households among some immigrant groups (23), and the subsequent high rates of Covid-19 infection among these groups (26), suggests that the housing problems should be addressed as an important social determinant of health, particularly in the case of Covid-19 among immigrants in Norway.

Other socio-economic barriers include first-line jobs, such as the transport sector, cleaning, health care, construction, etc. Participants reported that these jobs require commuting, despite the use of public transport being discouraged due to the Covid-19 pandemic in Norway. A previous report stated that immigrants in Norway mostly work in jobs that can be referred to as close-contact professions, and in which a home office is not an option (27). As most of these professions required close contact with people, immigrant workers may be easily exposed to Covid-19 infection, and the subsequent secondary spread to the community. Despite this, a recent study reported that occupation is not an important driver of the high rates of Covid-19 among immigrants from Somalia, Pakistan, Iraq, Afghanistan and Turkey (28). The study compared immigrants who hold several professions that require close contact with other people, including people with the same country of birth. However, participants in our study reported that people who work in close-contact professions serve as a vector of the infection in their community. Therefore, participants emphasized the secondary spread of infections from first-line workers to the families and the community at large. In line with our findings, a prior study stated that individual’s risk of infection depends on household size and occupation (25). Even so, we need to know more about the role of community characteristics and neighborhood factors on the uptake of preventive measures on Covid-19 among immigrants.

While socio-economic factors serve as a barrier to adherence, participants also stated socio-cultural factors that hinder them from adhering to preventive measures. Among these is the collectivist tradition. To combat the transmission, people have to avoid contact with others and to practice social distancing, which is very much contrary to how some immigrant groups operate. The number of Covid-19 outbreaks among immigrants in Norway was attributed to religious services and other private events. The collective tradition assumes that anyone in the community should participate in the community’s events, and that not complying may result in some form of social sanction. A prior study shows that 80% of infection in two provinces in South Africa resulted from church services and burial events as people resisted social distancing rules (29). Among some immigrant groups, the perceived obligation to social expectations, such as attending a wedding ceremony, burial or other community gathering may be stronger than the fear of contracting Covid-19. It is natural that immigrants from collectivist cultures often rely on support networks within communities of fellow immigrants, particularly those from common cultural backgrounds. For that, it is an incentive for every member to be part of the community, which is often proven by attending important community events. This tradition was repeatedly mentioned by study participants as a barrier to adherence to preventive measures among immigrants in Norway.

The authoritarian system at the immigrant’s country of origin has affected their compliance to governmental instructions. The reaction of immigrants to the Covid-19 pandemic may depend on certain features of their country of origin’s political culture, such as individuals’ compliance with collective habits, sense of civic duty, public trust and respect for governmental decisions. Almost all of the immigrant groups interviewed came from authoritarian countries where governments use strict measures, including arrests, to help enforce Covid-19 measures. According to frames of reference theory, immigrants operate with “multiple frames of reference,” including the home frame of reference (30). Given the “frame of reference effect,” immigrants may expect strict enforcement of mitigation measures from Norwegian authorities by means of arrest or fine, which is not the case in Norway. As immigrants’ expectations were not met, some immigrant individuals might have ignored complying with preventive measures. Nevertheless, the prior study among immigrants in Europe stated that “the ‘frame of reference effect’ weakens over time, and with increased acculturation in the country of residence” (31). We hypothesize that an increased integration of immigrants may decrease their attachment toward their countries of origin and the subsequent “frame of reference effect.”

Other barriers reported by this study include fear of stigma. Stigmatization is a social process designed to exclude those who are perceived to be a potential source of disease and may pose a threat to effective social living in the society (32). According to the participants, immigrants have been scapegoated for the spread of the virus in Norway. Unfortunately, immigrants subjected to stigma may also fuel harmful stereotypes, which may contribute to a situation that increases virus transmission as an immigrant may not only hide their illness to avoid discrimination, but that the stigma may also prevent them from adhering to mitigation measures, in addition to seeking health care. There is a long-established pattern of linking immigrants to disease, and stigmatizing them as the origin of a wide variety of diseases (33). Emphasizing the importance of communications to reduce stigma, the Norwegian Directorate of Migration (IMDI) released a call entitled, “Extraordinary grant for the provision of Covid-19 information to immigrants under the auspices of a voluntary organization.” The idea was to provide tailored information to immigrants, including reducing stigma and judgement to the people infected by the virus. Addressing the stigma experienced by immigrants during the Covid-19 era requires collaborative efforts from various stakeholders and requires all stakeholders to work together to ensure that nobody is stigmatized because of the pandemic.

The study has several limitations, and thus the results should be interpreted with caution. The study has used a qualitative design, with the results based on viewpoints of 88 purposely selected participants; therefore, the results cannot be generalized. However, we argue that the insight provided by the participants is of high relevance in providing tailored services to different immigrant groups, and for future research. Furthermore, participants were partially recruited from a resource group that serves as a breach between health institutions and immigrants, and who provide Covid-19 information to their respective communities in their local languages. Consequently, the information they provided may be partially influenced by their frustration with community members who do not listen and act upon the information provided. However, the interviews were conducted by researchers with a strong competence on migration and health, as well as prior experience with qualitative interviews. Therefore, we are confident that the data were collected professionally, with limited participant- and researcher-related bias.

Despite the limitations, the research described in this paper has two main implications. First, the study found that socio-economic factors such as overcrowding households negatively affect an immigrant’s ability to adhere to Covid-19 preventive measures. Overcrowding is a risk factor for various types of ill health, including both somatic and mental health. To reduce the inequality that arises from overcrowded housing, there is a need for a long-term strategy to improve the housing situation of low-income immigrant families who live in overcrowded households. The second implication is that there are also socio-cultural barriers that require special attention from health policymakers and government institutions that work for the health and integration of immigrants. The effect of socio-cultural barriers can be minimized through tailored information about Covid-19 among immigrants, as well as improved integration. Both socio-economic and socio-cultural factors reported by this study may not be limited to Covid-19, but may also influence the overrepresentation of immigrants in other illnesses, hence creating an avoidable inequality in health. Therefore, policymakers and health authorities should consider tailored policies and interventions that help improve the social determinants of health; only then can the immigrant’s health situation improve. The coronavirus pandemic should be taken as an alarm bell for the social gradient that has been creating health disparities among immigrants, not only during Covid-19, but also before the pandemic.

## Data Availability

All data produced in the present work are contained in the manuscript

## Data Availability Statement

The raw data supporting the findings of this paper will be made available on demand by the authors, without undue reservation.

## Ethics Statement

Ethical review and approval were not required for this study in accordance with the local legislation and institutional requirements. The participants provided their oral informed consent to participate in this study.

## Author Contributions

AG wrote the proposal and designed the study. NS & AG carried out the data analysis, interpretation, and drafted the manuscript. SQ and KP contributed the analysis and the write up of the manuscript. All authors contributed to the article and approved the submitted version.

## Acknowledgments

We would like to acknowledge Thor Indseth for securing the fund for this study. Thanks Charlott Nordstrøm, Hodon Dualeh, Selam Tsige, Mohamed Gawad, Ragnhild Spilker and Opinion company for collecting the data.

